# Identifying and Characterizing Bias at Scale in Clinical Notes Using Large Language Models

**DOI:** 10.1101/2024.10.24.24316073

**Authors:** Donald U. Apakama, Kim-Anh-Nhi Nguyen, Daphnee Hyppolite, Shelly Soffer, Aya Mudrik, Emilia Ling, Akini Moses, Ivanka Temnycky, Allison Glasser, Rebecca Anderson, Prathamesh Parchure, Evajoyce Woullard, Masoud Edalati, Lili Chan, Clair Kronk, Robert Freeman, Arash Kia, Prem Timsina, Matthew A. Levin, Rohan Khera, Patricia Kovatch, Alexander W. Charney, Brendan G. Carr, Lynne D. Richardson, Carol R. Horowitz, Eyal Klang, Girish N. Nadkarni

## Abstract

**Importance:** Discriminatory language in clinical documentation impacts patient care and reinforces systemic biases. Scalable tools to detect and mitigate this are needed.

**Objective:** Determine utility of a frontier large language model (GPT-4) in identifying and categorizing biased language and evaluate its suggestions for debiasing.

**Design:** Cross-sectional study analyzing emergency department (ED) notes from the Mount Sinai Health System (MSHS) and discharge notes from MIMIC-IV.

**Setting:** MSHS, a large urban healthcare system, and MIMIC-IV, a public dataset.

**Participants:** We randomly selected 50,000 ED medical and nursing notes from 230,967 MSHS 2023 adult ED visiting patients, and 500 randomly selected discharge notes from 145,915 patients in MIMIC-IV database. One note was selected for each unique patient.

**Main Outcomes and Measures:** Primary measure was accuracy of detection and categorization (discrediting, stigmatizing/labeling, judgmental, and stereotyping) of bias compared to human review. Secondary measures were proportion of patients with any bias, differences in the prevalence of bias across demographic and socioeconomic subgroups, and provider ratings of effectiveness of GPT-4’s debiasing language.

**Results:** Bias was detected in 6.5% of MSHS and 7.4% of MIMIC-IV notes. Compared to manual review, GPT-4 had sensitivity of 95%, specificity of 86%, positive predictive value of 84% and negative predictive value of 96% for bias detection. Stigmatizing/labeling (3.4%), judgmental (3.2%), and discrediting (4.0%) biases were most prevalent. There was higher bias in Black patients (8.3%), transgender individuals (15.7% for trans-female, 16.7% for trans-male), and undomiciled individuals (27%). Patients with non-commercial insurance, particularly Medicaid, also had higher bias (8.9%). Higher bias was also seen in health-related characteristics like frequent healthcare utilization (21% for >100 visits) and substance use disorders (32.2%). Physician-authored notes showed higher bias than nursing notes (9.4% vs. 4.2%, p < 0.001). GPT-4’s suggested revisions were rated highly effective by physicians, with an average improvement score of 9.6/10 in reducing bias.

**Conclusions and Relevance:** A frontier LLM effectively identified biased language, without further training, showing utility as a scalable fairness tool. High bias prevalence linked to certain patient characteristics underscores the need for targeted interventions. Integrating AI to facilitate unbiased documentation could significantly impact clinical practice and health outcomes.

## INTRODUCTION

Discriminatory language in clinical documentation causes enduring harm by perpetuating biases and contributing to worse patient outcomes.(1) (2) Such language infiltrates electronic health records (EHRs) through providers’ implicit biases and subjective assessments.(3) One particularly damaging form of bias is the use of “negative descriptors” like “aggressive” or “resistant”, which stigmatize patients, especially those from marginalized groups.(4) (5) (6) Significant disparities exist in the application of these terms, particularly against Black patients, which may reflect underlying biases in clinical documentation.(3) Furthermore, this biased language may exacerbate racial inequities and reduce patient trust, especially as patients gain increased access to their EHRs under the 21st Century Cures Act.(7)

Understanding the impact of biased language in medical records requires investigation into underlying factors, including patient demographics and provider characteristics.(8) (9) Factors such as racism and stigmatized diagnoses can amplify the effects of bias, underscoring the need for interventions.(5) (6) (7) (8) (9) (10) Categorizing bias allows for the identification of specific patterns, such as how different types of bias interact and compound each other. Previous research has been limited by its narrow focus, often addressing bias within medical documentation on a case-by-case basis or within single-axis frameworks like race or gender, as well as by the labor-intensive nature of manual chart review.(8) (9) This highlights the need for scalable tools to detect and categorize bias within EHRs.

Large language models (LLMs) offer a promising solution by detecting negative descriptors in EHRs through generalization from existing data, bypassing the need for specific training.(11) Prompt engineering further refines this process, enabling LLMs to identify and categorize bias patterns within clinical notes consistently and at scale.(11) (12) (13)

The adaptability of LLMs across various healthcare systems and EHR formats is crucial for tracking and comparing linguistic features that may indicate bias. Ultimately, this surveillance can drive systemic improvements by embedding bias awareness into everyday clinical practice, standardizing documentation across healthcare settings, and leveraging technology to consistently identify and correct biased language.(14) By doing so, they foster a culture of equity and respect, enhance the accuracy and neutrality of patient records, and ultimately lead to more equitable treatment outcomes across diverse groups.

The Emergency Department (ED) presents a unique opportunity for investigating explicit bias in healthcare documentation. The ED, as the initial patient contact point, encounters diverse medical conditions and demographics, making it critical for studying bias in clinical judgment, documentation, and its downstream influences on clinical care.(15) (16) Inpatient settings, where precise and unbiased documentation is necessary for ongoing clinical care, present a different set of challenges and opportunities.(17) (18)

We aimed to leverage the capabilities of GPT-4 to identify and categorize bias in EHRs from ED visits and hospital discharge summaries. We also evaluated provider ratings and the acceptability of alternative debiasing language suggested by LLMs. Finally, we make the prompts and documentation available for detecting and mitigating bias in clinical documentation.

## METHODS

### Study Framework

The Mount Sinai Health System (MSHS) institution-wide, multidisciplinary Factual, Affirming, Informative, and Respectful (FAIR) Documentation Workgroup (WG) was formed in June 2022 to enhance health equity by addressing biased language. FAIR employed a phased approach to identify and analyze negative descriptors in documentation. Initial phases involved stakeholder collaboration, literature review, developing a term library, and reviewer training. This study was developed as part of the FAIR WG initiative and introduced an AI-assisted methodology using GPT-4. **Figure 1** provides an overview of the transition from FAIR’s human-centered approach to an AI-assisted methodology. This study received approval from the Institutional Review Board (IRB) of the Icahn School of Medicine at Mount Sinai.

**Figure 1.**
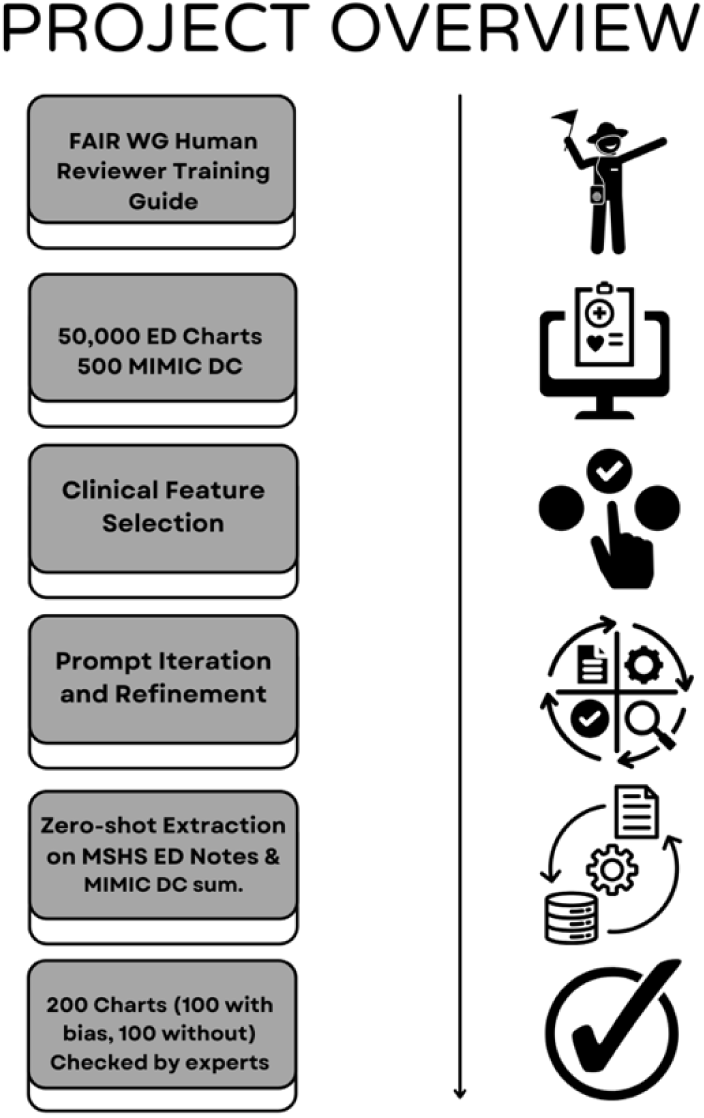
Overall Study Design and Methodology.

### Study Population

We used 50,000 Adult MSHS ED notes from 2023 and 500 Medical Information Mart for Intensive Care (MIMIC-IV) discharge summaries.(19) One note was randomly selected for each unique patient. Further details are described in the supplementary appendix.

### Prompt Refinement and Zero-Shot Extraction Process

We engineered the prompts for zero-shot extraction. GPT-4 was assigned the role of an emergency physician specialized in recognizing biased documentation. It was provided with definitions of bias categories developed for human reviewers. We evaluated GPT-4 for identifying and categorizing bias and suggesting unbiased alternatives. We made iterative refinements by reviewing GPT-4 performance on 100 EHR sentences, split evenly between biased and unbiased text. These examples were solely used for prompt optimization and did not involve any fine-tuning or training of the model itself. For final prompt text see **Supplementary Table 1**.

We used the GPT-4 application programming interface (API) to identify and categorize bias in clinical notes, prompting it to flag bias and its subtypes. API responses in JSON format were parsed to tabulate biases. In 0.2% of cases where API calls failed due to OpenAI’s GPT deployed in Azure compliance rules, two emergency providers with expertise in documentation bias (AM and IT) manually analyzed notes for bias presence and type. Manually analyzed cases were excluded from the primary GPT-4 and manual review comparison to ensure unbiased validation.

### Manual Validation

To validate GPT-4 output, 200 MSHS and 50 MIMIC-IV charts were randomly selected, evenly split between “no bias” and “with bias” labels. Reviewers (AM and IT) independently evaluated the output for accuracy, resolving disagreements through discussion (**Supplementary Figure 1**). They also assessed GPT-4’s language improvement suggestions on a 10-point scale (Scores: 0 = Much worse, 5 = No change or similar, 10 = Much Improved) (**Supplementary Table 2**).

### Modifying Feature Selection

The approach to identifying clinical features that modify negative descriptors presence was informed by literature review on patient characteristics and provider characteristics associated with bias (**Supplementary Table 3**).(20) (21) (22)

### Statistical Analysis

To validate prompt outputs, we compared them to assessments from human reviewers with high inter-rater reliability (IRR). We quantified concordance using Cohen’s kappa, Fleiss’ kappa, and percentage agreement. The prompt underwent three rounds of iteration until GPT consistently performed comparably to human reviewers (IRR ≥ .65), (**Supplementary Figure 2**). We then applied the final prompt to MSHS ED notes and MIMIC-IV discharge summaries. GPT-4 was then utilized to flag potential biases in clinical notes. For the MSHS cohort, we conducted a univariate analysis using chi-square tests to examine associations between bias flags and various features. Analyses were performed using Python 3.9 between February and May 2024.

## RESULTS

### Dataset for ED Visits

We analyzed 424,427 ED visits from 2023. After applying exclusion criteria, 230,967 unique patient records remained. We selected a random sample of 50,000 unique patients, one visit per patient, each represented by a single, detailed note (> five words). **Table 1** displays descriptive statistics of the study population, including demographics and socioeconomic factors (see **Supplementary Table 4** for complete descriptive analysis).

**Table 1.** Demographic Characteristics and Counts for MSHS ED Included Population.

|  |  |  |
| --- | --- | --- |
| **Feature\*** | **Category** | **Number (%)** |
| Age Group | 18-39 | 16,894 (33.8) |
|  | 40-59 | 14,552 (29.1) |
|  | 60-79 | 13,485 (27.0) |
|  | 80+ | 5,069 (10.1) |
| Race | Black | 15,711 (31.4) |
|  | Asian | 2,536 (5.1) |
|  | White | 12,673 (25.3) |
|  | Unknown/Other | 19,080 (38.2) |
| Ethnicity | Hispanic | 14,950 (29.9) |
|  | Unknown/Other | 35,050 (70.1) |
| Sex | Female | 27,019 (54.0) |
|  | Male | 22,977 (46.0) |
| Gender Identity | Female | 14,646 (29.3) |
|  | Male | 11,384 (22.8) |
|  | Non-Conforming | 116 (0.2) |
|  | Trans-Female | 70 (0.1) |
|  | Trans-Male | 18 (0.0) |
|  | Not Disclosed | 23,766 (47.5) |
| Sexual Orientation | Straight | 8,953 (17.9) |
|  | Bisexual | 307 (0.6) |
|  | Gay or Lesbian | 744 (1.5) |
|  | Unknown | 39,996 (80.0) |
| Marital Status | Single | 28,788 (57.6) |
|  | Married | 11,378 (22.8) |
|  | Widowed | 1,907 (3.8) |
|  | Separated | 3,613 (7.2) |
|  | Divorced | 1,684 (3.4) |
|  | Unknown/Other | 2,630 (5.3) |
| Religion | Christian | 22,452 (44.9) |
|  | Muslim | 2,246 (4.5) |
|  | Jewish | 1,996 (4.0) |
|  | Buddhist | 205 (0.4) |
|  | Hindu | 201 (0.4) |
|  | Unspecified | 5,260 (10.5) |
|  | Other | 4,597 (9.2) |
| Preferred Language | English | 43,729 (87.5) |
|  | Spanish | 4,581 (9.2) |
|  | Not Collected | 204 (0.4) |
|  | Other | 1,486 (3.0) |
| County | New York | 23,932 (47.9) |
|  | Bronx | 4,134 (8.3) |
|  | Kings | 7,721 (15.4) |
|  | Queens | 9,206 (18.4) |
|  | Richmond | 263 (0.5) |
|  | Brooklyn | 169 (0.3) |
|  | Other | 4,575 (9.2) |
| Housing Status | Domiciled | 47,697 (95.4) |
|  | Undomiciled | 2,303 (4.6) |
| Payor Financial Class | Medicaid | 18,531 (37.1) |
|  | Medicare | 14,618 (29.2) |
|  | Commercial/Managed | 12,787 (25.6) |
|  | Other | 4,064 (8.1) |
| *Features extracted from MSHS Epic EHR, primarily from structured fields. Housing status indirectly classified through clinical documentation. Race/ethnicity based on Mount Sinai's standardized mappings. Gender identity and sexual orientation were self-reported. Top 20 chief complaints included. |  |  |

The study population was diverse. Age distribution was relatively balanced across adult groups, with a slight majority (62.9%) under 60 years old. Racially, Black patients represented 31.4%, White patients 25.3%, and a substantial 38.2% in the Unknown/Other category. Ethnically, 29.9% identified as Hispanic. Sex distribution slightly favored females (54.0% vs 46.0% males). For gender identity, a notable 47.5% did not disclose their gender. Most patients were single (57.6%) and preferred English (87.5%). Geographically, patients were predominantly from New York County (47.9%). Socioeconomically, 95.4% were domiciled, and Medicaid was the most common payer (37.1%)

### Overall Description of Bias at MSHS

Analysis of 50,000 MSHS ED notes revealed bias in 3,229 (6.5%) notes. Broken down into different types of bias, discrediting language was seen in 1,989 (4.0%), stigmatizing/labeling in 1,678 (3.4%), judgmental language in 1,593 (3.2%), and stereotyping observed in only 281 (0.6%) notes (**Supplementary Figure 3**).

### Performance Metrics by Bias Type At MSHS

In terms of diagnostic test performance, GPT showed strong sensitivity and specificity across different bias types. The sensitivity for identifying “Any Bias Present” was 95.5%, while the specificity was 85.7%. Similarly, the sensitivity for “Judgmental” bias was 97.7%, with a specificity of 94.3%. Positive Predictive Values (PPV) and Negative Predictive Values (NPV) also indicated robust performance, with PPVs ranging from 42.9% for “Stereotyping” to 82.4% for “Judgmental”, and NPVs consistently high, such as 98.4% for “Stereotyping” and 99.3% for “Judgmental” (**Table 2**).

**Table 2.** Performance Metrics for GPT EHR Bias Detection.

| **Table 2.** Performance Metrics for GPT EHR Bias Detection |  |  |  |  |
| --- | --- | --- | --- | --- |
| **Bias Type** | **Sensitivity** | **Specificity** | **PPV** | **NPV** |
| Any Bias Present | 0.95 | 0.86 | 0.84 | 0.96 |
| Stigmatizing/Labeling | 0.93 | 0.94 | 0.80 | 0.98 |
| Judgmental | 0.98 | 0.94 | 0.82 | 0.99 |
| Discrediting | 0.93 | 0.90 | 0.77 | 0.97 |
| Stereotyping | 0.50 | 0.98 | 0.43 | 0.98 |

### Examples of GPT-4 Flagged Documentation Bias at MSHS

We reviewed flagged charts for four bias types (**Table 3**): discrediting (language undermining patient credibility, e.g., “patient claimed”), judgmental (terms imposing moral judgment, e.g., “snorting and injecting heroin to cope with pain”), stereotyping (assumptions based on demographics, e.g., undomiciled patients don’t have real medical complaints), and stigmatizing/labeling (negative patient labels, e.g., “disrespectful yelling and cursing”).

**Table 3.** Examples of Negative Descriptors Categories in the EHR.

| **Table 3.** Examples of Negative Descriptors Categories in the EHR |  |  |
| --- | --- | --- |
| **Example of Discrediting Bias in EHR** |  |  |
| Clinical Note: Unwitnessed fall but patient claimed "he tripped and fell" accompanied by HHA OOB with assistance only. At baseline with laceration at the back of the head. |  |  |
| Bias Term | Bias Type | GPT Reasoning |
| Patient claimed | Discrediting | 'Claimed' could be interpreted as the physician questioning the legitimacy of the patient's report of how the fall occurred. |
| **Example of Judgmental Bias in EHR** |  |  |
| Clinical Note: Patient states he fell and injured his hip last week and has been snorting and injecting heroin to cope with pain. Last heroin use was yesterday. |  |  |
| Bias Term | Bias Type | GPT Reasoning |
| Snorting and injecting heroin to cope with pain | Judgmental | Suggests a moral judgment against the patient's method of dealing with pain. |
| **Example of Stereotyping Bias in EHR** |  |  |
| Clinical Note: 62M w/hx Afib (on eliquis), CVA, hyperthyroidism, etoh use disorder, htn, BIBEMS after someone called 911 as he was slumped on bench... Pt sleeping, opens eyes to verbal stimuli but turns away from provider and asks to be left alone.... No signs of head trauma, MAE. C/f acute alcohol intoxication vs. Shelter seeking behavior. |  |  |
| Bias Term | Bias Type | GPT Reasoning |
| C/f acute alcohol intoxication vs shelter seeking behavior | Stereotyping | Generalizes and assumes behavior based on the patient's history of alcohol use disorder, implying his visit is either due to intoxication or to seek shelter rather than to seek medical attention. |
| **Example of Stigmatizing/Labeling Bias in EHR** |  |  |
| Clinical Note: Patient is c/o pain. Pain meds is given as order... patient is being disrespectful yelling and cursing and stated we didn't know what we were doing and wanted to report told patient to go ahead he has that right to do so. |  |  |
| Bias Term | Bias Type | GPT Reasoning |
| Patient is being disrespectful yelling and cursing | Stigmatizing/Labeling | Labels the patient as 'disrespectful', 'yelling' and 'cursing', reducing them to their behavior. It contributes to stigma and depersonalizes the individual. |

### Scores for GPT-4 Suggestions of Non-Biased Language at MSHS

Reviewers rated GPT-4’s language suggestions on a 10-point scale. Average scores across bias categories were Stigmatizing/Labeling 9.47, Judgmental 9.83, Discrediting 9.61. Full analysis and examples of original text with GPT’s unbiased suggestions can be found in **Supplementary Tables 2 & 5**.

### Bias by Relevant Clinical Features

The univariate analysis revealed significant bias variations across demographic, socioeconomic, and health-related factors in clinical documentation.

#### Demographic Disparities

Patients with non-conforming gender identities faced significantly higher bias rates compared to those who did not identify as non-conforming (14.7-16.7% vs. 4.8-8.5%, p<0.001). This group includes primarily cisgender individuals but may also include others who do not identify as non-conforming. Bisexual patients experienced elevated bias compared to heterosexual patients (9.4% vs. 5.7%, p<0.001). Black patients encountered higher bias rates than White or Asian patients (8.3% vs. 6.0% and 3.4% respectively, p<0.001). Notably, Hispanic ethnicity and non-English language preference did not correlate with increased bias.

#### Socioeconomic Influences

Housing status emerged as a critical factor, with undomiciled patients facing over five times the bias rate of domiciled patients (27.0% vs. 5.5%, OR 6.37, 95% CI 5.74-7.07, p<0.001). Medicaid recipients experienced higher bias compared to those with commercial insurance (8.9% vs. 3.3%, p<0.001), highlighting potential disparities in care based on insurance type.

#### Health-Related Characteristics

Frequent ED utilization strongly correlated with increased bias, with patients having over 100 previous visits (all time) experiencing four times the bias rate of those with 0-3 visits (21.0% vs. 5.2%, p<0.001). Active smokers faced significantly higher bias compared to non-smokers (13.8% vs. 4.2%, p<0.001), suggesting potential stigma associated with this health behavior.

#### Clinical Encounter Details

Chief complaints related to substance use disorder and psychiatric conditions exhibited exceptionally high bias rates (32.2% and 25.0% respectively), far exceeding those for other medical conditions (e.g., 2.5% for general complaints, p<0.001). Physician-authored notes showed higher bias rates than those by other providers (9.4% vs. 5.2% for nurses, p<0.001), and overnight shifts were associated with increased bias (9.6% vs. 5.3% for day shifts, p<0.001). **Table 4** summarizes key findings, revealing significant disparities across various patient characteristics and clinical factors (p<0.001 for all listed features).

**Table 4.**
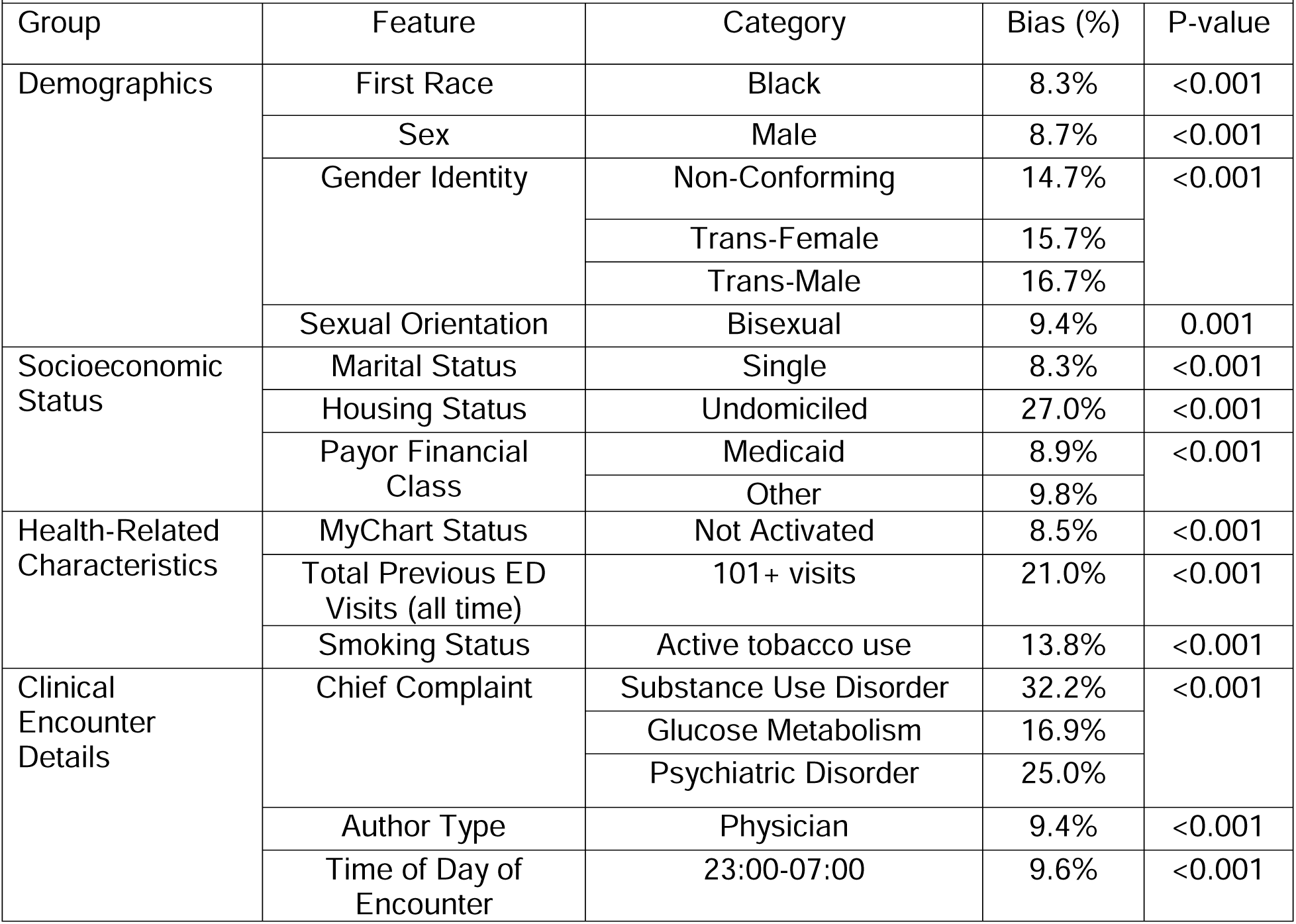
Key Results of Univariate Analysis of Overall Bias in ED Notes.

| **Table 4.** Key Results of Univariate Analysis of Overall Bias in ED Notes |  |  |  |  |
| --- | --- | --- | --- | --- |
| Group | Feature | Category | Bias (%) | P-value |
| Demographics | First Race | Black | 8.3% | <0.001 |
|  | Sex | Male | 8.7% | <0.001 |
|  | Gender Identity | Non-Conforming | 14.7% | <0.001 |
|  |  | Trans-Female | 15.7% |  |
|  |  | Trans-Male | 16.7% |  |
|  | Sexual Orientation | Bisexual | 9.4% | 0.001 |
| Socioeconomic Status | Marital Status | Single | 8.3% | <0.001 |
|  | Housing Status | Undomiciled | 27.0% | <0.001 |
|  | Payor Financial Class | Medicaid | 8.9% | <0.001 |
|  |  | Other | 9.8% |  |
| Health-Related Characteristics | MyChart Status | Not Activated | 8.5% | <0.001 |
|  | Total Previous ED Visits (all time) | 101+ visits | 21.0% | <0.001 |
|  | Smoking Status | Active tobacco use | 13.8% | <0.001 |
| Clinical Encounter Details | Chief Complaint | Substance Use Disorder | 32.2% | <0.001 |
|  |  | Glucose Metabolism | 16.9% |  |
|  |  | Psychiatric Disorder | 25.0% |  |
|  | Author Type | Physician | 9.4% | <0.001 |
|  | Time of Day of Encounter | 23:00-07:00 | 9.6% | <0.001 |

### Bias Subtypes

The complete univariate analyses of bias subtypes with confidence intervals is available in **Supplementary Table 6.**

#### Stigmatizing/Labeling

Undomiciled patients and those with over 100 ED visits experienced particularly high bias (16.5%, 14.2%, respectively). Clinical conditions also played a significant role, with substance use disorder and psychiatric conditions showing elevated bias rates (24.6%, 15.5%, respectively).

#### Judgmental

Undomiciled patients and those with over 100 ED visits encountered increased bias (14.2%, 9.8%, respectively). Patience with substance use disorder (14.5%), glucose metabolism disorders (14%), and individuals identifying as bisexual (6.8%) experienced notable rates.

#### Discrediting

Undomiciled individuals faced a 16.9% bias rate, while those with over 100 ED visits experienced a 12.7% bias rate. Substance use disorder stood out with a 19.2% bias rate, aligning with broader societal stigma surrounding this condition.(23)

#### Stereotyping

While uncommon overall (0.6% across all patients), undomiciled patients and trans-male individuals faced substantial stereotyping with bias rates of 5.0% and 5.6%, respectively. These rates were notably higher than the average, suggesting persistent stereotyping for these marginalized populations.(24)

### Bias in MIMIC-IV

500 discharge summaries from the MIMIC-IV dataset revealed an overall bias of 7.4%, compared with 6.5% from the MSHS dataset. The MIMIC-IV notes showed increased stigmatizing/labeling (5.4%), judgmental language (5.8%), and discrediting language (5.0%), with lower stereotyping at 0.6%, compared to the MSHS dataset (**Supplementary Figure 4**).

## DISCUSSION

We evaluated zero-shot learning with GPT-4 for identifying and mitigating bias in clinical documentation. Previous studies identified bias in clinical documentation,(4) (5) but these approaches have not been scalable. We advance the field by using a frontier LLM for automated bias detection and categorization. Our findings include the following: (1) The prevalence of bias in ED notes was 6.5%, and in discharge notes, 7.4%, with LLM bias detection being accurate compared to manual review; (2) Bias was more prevalent with key clinical features, notably housing instability, frequent ED visits, and substance use disorder; (3) Suggested debiasing revisions by the LLM were rated highly by physicians.

This scalable approach addresses a significant gap in large EHR dataset bias detection.(25) As patients increasingly access their EHRs, biased language critically impacts patient-provider relationships and trust. Differences in bias rates and types between healthcare settings suggest that documentation practices and institutional cultures influence bias prevalence.

Our analysis revealed disparate rates of bias for certain groups, including transgender individuals, Black patients, and those with socioeconomic challenges or substance use disorders. The disparity, particularly in mental health and substance use documentation, potentially reflects and reinforces societal stigmas.(26)

Biased language reflects implicit bias in the clinical encounter and suggests that structural and societal biases are being codified into the EHR.(26) (27) Our findings underscore the need for a comprehensive approach to bias mitigation, including targeted education and systemic changes in documentation practices.(28) (29)

GPT-4 excelled in debiasing language, receiving high scores from human reviewers, particularly for judgmental and stigmatizing/labeling suggestions. Nevertheless, the discrepancies between Cohen’s Kappa and percentage agreement, particularly for “Stereotyping,” highlight the challenges in consistently identifying subtle forms of bias. This underscores the need for ongoing refinement of AI tools, especially in capturing nuances.

It is important to acknowledge that LLMs like GPT-4 may themselves harbor inherent biases derived from their training data. These models are trained on vast amounts of text that can contain societal prejudices and stereotypes, potentially influencing the model’s outputs.(30) (31) (32) (33) In the context of detecting clinical bias, the LLM might inadvertently reflect these biases, leading to inconsistent identification or introducing new biases through its suggestions. Additionally, phenomena such as sycophancy bias—where models align their responses with presumed user expectations— can impact the LLM’s performance.(30)

While LLMs like GPT-4 can assist in identifying biased language, probing their outputs by evaluating metacognition is crucial.(34) The subjective nature of bias detection means that what one clinician perceives as bias, another might interpret as appropriate clinical practice. Therefore, bias detection should be performed within a sociotechnical framework that combines technological tools with human expertise.(35) Such an approach acknowledges that technology alone cannot fully capture the nuances of bias in healthcare.(36) Successful examples of sociotechnical systems in healthcare demonstrate that integrating AI with human oversight can enhance outcomes while mitigating risks associated with algorithmic bias.(37)

Additionally, it is crucial to clarify that the intent of these suggestions is not to directly alter the content of clinical notes. Simply changing the language does not address the underlying biases. Instead, the goal is to expose clinicians to a variety of unbiased language options, encouraging critical reflection on their language choices and the broader implications for patient care. By incorporating LLMs as supportive tools rather than definitive arbiters, we can promote critical reflection among clinicians on their language choices and the broader implications for patient care. Integrating AI-suggested language into medical education and residency training could enhance cultural competency, aligning with calls for more comprehensive approaches to providing equitable healthcare.(38) (39) By doing this, we can foster critical reflection on language choices in patient care.(40) (41)

The implications of these findings for health systems are significant. AI approaches could serve as powerful screening tools for bias in clinical documentation, enabling comprehensive surveys of existing records to identify patterns and trends in biased language use.(42) (43) This approach allows health systems to strategically address documentation bias at a systemic level. AI-generated reports could highlight areas of concern, such as higher rates of stigmatizing language in ED notes or discharge summaries.(13) (25) (42) These insights could inform educational interventions, such as workshops on inclusive language for high-risk departments or the development of specialty-specific documentation guidelines.(22) Over time, these reports could track progress, allowing administrators to assess the effectiveness of interventions and adjust strategies accordingly.

This study has several limitations that warrant consideration. First, a substantial amount of missing or undisclosed data across key demographic variables, particularly in race, ethnicity, gender identity, and sexual orientation, may lead to incomplete conclusions. The small sample sizes for non-binary gender identities and non-heterosexual orientations further limit our ability to draw robust conclusions about these groups.

Second, bias in healthcare is multifaceted, shaped by various factors, including patient characteristics, physician characteristics, the nature of the medical complaint. while we focused on patient characteristics and timing of care, we did not fully account for other potential confounders, such as patient age—which may contribute to potential ageism— English proficiency, type of complaint, time of day, and day of the week. Incorporating these variables would further strengthen the analysis and help disentangle this very complex construct of bias in healthcare.

Third, GPT-4 inherently carries biases from its training, which can reflect and perpetuate societal prejudices, particularly those related to demographics, disease prevalence, and treatment outcomes.(30) (31) Our human validation process helps mitigate this concern, but two-reviewer human validation may not always work. The subjective nature of bias definitions reflects the complexity of clinical communication; a model trained at one-time point, and with specific training, might not be generalizable. Our high IRR does, however, suggest a robust categorization approach. Additionally, some terms deeply embedded in EHRs, such as “complains” in “chief complaint,” were not flagged in this study, though they may still carry inherent biases.

Fourth, we did not assess the severity or magnitude of bias within each category or account for the possibility of multiple bias infractions, of the same category, occurring within a single note. This approach likely underestimates both the frequency and overall impact of biased language in clinical documentation.

Fifth, the perception of certain terms can vary based on patient or provider demographics; for example, “queer” may be seen as negative by older individuals but reclaimed positively by younger generations.(44) We did not conduct a diachronic linguistic analysis, which considers how word meanings change over time. This limitation is particularly relevant given the different timeframes of the MSHS and MIMIC-IV datasets, where the meaning of certain terms may have shifted. However, this issue does not affect the contemporaneous comparisons between ChatGPT and human reviewers. The importance of accounting for temporal variations, should however, be accounted for in future studies.

Sixth, we did not examine the distribution of note writers, leaving unclear whether biased terms were concentrated among a few providers or spread across many. We also did not quantify the percentage of copy/pasted content in notes, which could potentially amplify the propagation of biased language.(37)

Seventh, our study’s focus on New York City boroughs and the overrepresentation of Medicaid and Medicare patients may limit generalizability to other settings or populations. Additionally, the predominance of English-speaking patients may not adequately capture the experiences of linguistic minorities. Furthermore, the lack of comprehensive socioeconomic indicators beyond housing status and payor class could mask important confounding variables.

Finally, our study was limited to univariate analyses, and a multivariate analysis could have provided more nuanced insights into the interplay of various factors contributing to biased documentation.

## CONCLUSIONS

This study advances understanding of clinical documentation bias using GPT-4 Zero-Shot Learning. GPT-4 effectively identified and categorized bias and suggested alternatives, offering a scalable approach to monitoring bias in EHR documentation. This AI-driven method enables strategic bias intervention, though ethical implementation requires stakeholder engagement and human oversight.

Future research should compare fine-tuned healthcare-specific AI models with general models like GPT-4, benchmark and mitigate biases in AI models and their training data, analyze provider-level patterns of biased language, investigate copy/paste practices, and conduct multivariate analyses. Crucially, researchers must examine how biased documentation affects care quality, patient perceptions, and engagement, and develop interventions.

These efforts will translate findings into actionable strategies, improving healthcare documentation and patient care. While this study provides a foundation for AI-assisted bias reduction, ongoing refinement and validation are essential for advancing equitable and effective healthcare delivery.

## Data Availability

All data produced in the present study are available upon reasonable request to the authors

