## SUPPLEMENTAL APPENDIX for "Identifying and Characterizing Bias at Scale in Clinical Notes Using Large Language Models"

**Study Population**

MSHS

The Mount Sinai Health System (MSHS) serves a diverse urban population in New York City, encompassing patients from various ethnic, socioeconomic, and cultural backgrounds, reflecting the city's heterogeneous demographic profile. Notes were collected over 12 months (2023), ensuring a broad representation of clinical activity and mitigating seasonal biases.

Medical Information Mart for Intensive Care, Version 4 (MIMC-IV)^1^

Extending the study beyond the MSHS ED, MIMIC-IV contains de-identified health data from 145,915 patients admitted to the Beth Israel Deaconess Medical Center in Boston, Massachusetts, between 2008 and 2019, representing a mix of urban and suburban populations with a broad range of medical conditions. MIMIC has been pivotal in uncovering bias in healthcare through previous studies, showcasing disparities in patient treatment and outcomes.^1-3^ Its established role in benchmarking machine learning tools in healthcare underscores its importance as a comparative resource, essential for identifying and addressing embedded biases in clinical algorithms.^4,5^

**Data Collection and Preprocessing**

The preprocessing and analysis were conducted in a secure, HIPAA-compliant environment using a specialized GPT-4 version 0613 on the Microsoft Azure MSHS tenant. This infrastructure provided robust security, scalability for the extensive dataset, and advanced computational capabilities, while integrating GPT-4's processing power with stringent privacy requirements to protect sensitive patient information.

**Data Extraction**

For MHSH, data collection was confined to unique patient visits with complete medical or nursing notes, excluding pediatric records, psychiatric evaluations, and repetitive entries. Features were extracted from MSHS Epic EHR using SQL-based extract, transform, and load (ETL) processes, mostly from structured fields. Housing status required indirect classification, comparing addresses to a shelter database and searching ED notes for homelessness indicators. Race and ethnicity followed standardized mappings from Mount Sinai's Diversity and Inclusion Office, which are derived from US Census classifications. Gender identity and sexual orientation were determined by self-reporting. Finally, the top 20 chief complaints, during the study duration, were included for feature analysis. For MIMC-IV, only one discharge summary per person was included, and modifying features were not assessed.

**Selection of GPT-4**

GPT-4, a model from the Generative Pre-trained Transformer series by OpenAI, was chosen for its advanced language understanding capabilities, essential for interpreting the complex structures in electronic medical records.^6^ Its extensive training allows it to effectively interpret medical terminologies and identify biases, leveraging zero-shot learning capabilities.^7,8^ Additionally, many health systems are already utilizing GPT-4 on Azure, which guided our decision to use it, ensuring compatibility and ease of adoption for other systems. We used GPT-4 version 0613 with default hyper-parameters.

| **Supplementary Table 1.** Final Prompt Framework and Instructions | |
| --- | --- |
| **Prompt Framework** | **Prompt Instructions** |
| **Role** | You are a highly esteemed emergency physician recognized for your expertise in identifying and addressing biased language in clinical documentation. |
| **Instructions** | Your mission is to scrutinize emergency department chart excerpts, focusing specifically on detecting language that may contribute to patient stigmatization. For this task, we categorize "stigmatizing language" into four distinct types: Discrediting, Judgmental, Stigmatizing/Labeling, and Stereotyping. |
| **Steps** | **Below are the definitions and guidelines for each category: Discrediting**: Language that unjustifiably questions the legitimacy of a patient's self-reported symptoms or concerns. Tag language as discrediting only if it shows an unjustified pattern of skepticism not supported by clinical evidence or dismisses patient reports without proper investigation. Specifically pay attention when adverbs such as reportedly, allegedly, and supposedly, and/or similar words like "claims", are used to modify the validity of what is being presented. |
|  | **Judgmental**: Language that conveys moral judgment or reflects personal values onto the patient. Identify language as judgmental only when it implies moral failing or character flaws unrelated to the clinical context. Specifically include the phrase "non-compliance" when given without reason, or "toxic habits", or "no toxic habits" or any other phrasing that implies the patient has failed in some manner. Ignore terms that are factual and have medical implications like "active smoker", "toxic appearing", or "ill appearing." |
|  | **Stigmatizing/Labeling**: Language that unfairly reduces a patient to their medical condition can be stigmatizing. Pay specific attention to when a patient is referred to as a negative adjective (patient is belligerent, agitated, drug-seeking behavior, and/or aggressive). Consider language stigmatizing when it unnecessarily labels patients with negative adjectives, contributing to stigma or depersonalizing the individual. Any references to medical history, age, and clinically relevant behaviors are not biased when they are stated factually and without negative adjectives that could imply judgment. Ignore medical conditions in this assessment (e.g. patient has a history of Alcoholic gastritis, other disorders, obesity, dementia, abuse disorders), unless the patient is being referred to as a medical condition (e.g. Patient is a "diabetic" or "sicker" or "asthmatic"). |
|  | **Stereotyping**: Presumptions about a patient based on demographic groups and without individual evidence. Mark language as stereotyping when it applies generalized traits or behaviors to a broad group rather than assessing the individual patient (pay specific attention to marginalized groups like racial/ethnic minorities, women, elderly, patients dealing with poverty, non-English speakers, patients dealing with homelessness, and patients who identify as LGBTQIA. For the purposes of this category only, ignore bias mentioned in past medical history, exam findings about mental status and alertness (as these tend to have clinical value). Also ignore when a patient is referred to as their age and gender. |
| **End Goal** | **Review each emergency department chart excerpt provided.** |
|  | Identify and categorize any instance of language that falls into the above categories. |
|  | For each identified instance, explain why it has been categorized as such, referencing the guidelines provided. |
|  | For each identified instance, provide a brief example revision, that demonstrates how the bias could be eliminated from the documentation. |
|  | Bias in clinical notes can fall into multiple categories, but please try to find the most pertinent fit for instance. Drug seeking behavior can be considered discrediting, judgmental, Stigmatizing/Labeling, and Stereotyping depending on the use, but it most falls in line with Stigmatizing/Labeling for this review as it usually refers to the patient as an adjective. |
|  | Objective: Your goal is to enhance the awareness and understanding of how language can influence patient care and perception in clinical settings. This exercise will contribute to developing best practices for creating unbiased and respectful clinical documentation. Keep your responses brief and to-the-point. |
| **Narrow** | Expected JSON Format for Response:  [{"bias term": "<biased phrase verbatim>", "bias type": "<type of bias - Discrediting, Judgmental, Stigmatizing/Labeling, Stereotyping>", "reason": "<short explanation why the phrase is considered biased>" "revision": "<suggest a concise revision which will not show bias>"},...]  If there are no bias terms in the note please return: "None" |

**Supplementary Figure 1.** GPT Vs. Expert Reviewers for Negative Descriptor Detection in MSHS Dataset

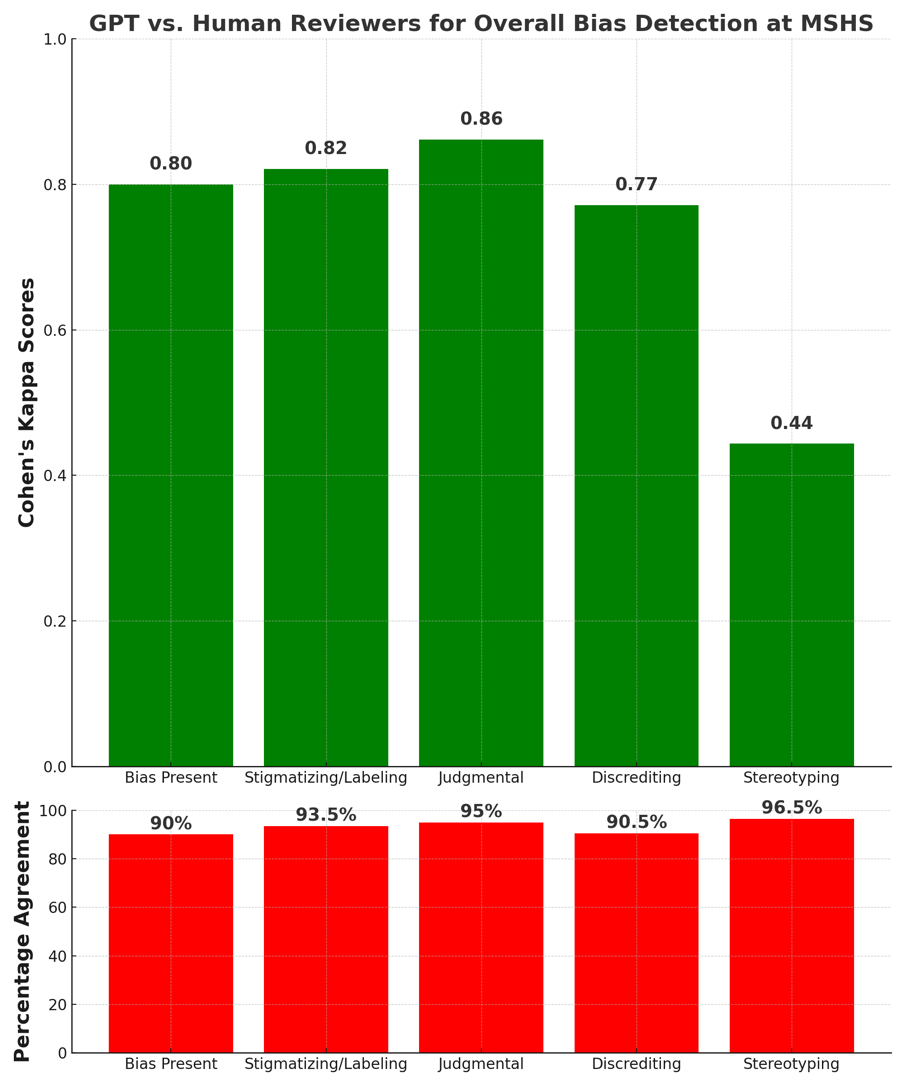

| **Supplementary Table 2.** Complete Scores for GPT-4 Suggestions of Unbiased Language at MSHS | | | |
| --- | --- | --- | --- |
| **Bias Category** | **Bias Present** | **Average Score of GPT Suggestions** | **Number of Observations** |
| Stigmatizing/Labeling | TRUE | 9.47 | 44 |
| Judgmental | TRUE | 9.83 | 43 |
| Discrediting | TRUE | 9.61 | 53 |
| Stereotyping | TRUE | 9.5 | 6 |
| Scores: 0 = Much worse, 5 = No change or similar, 10 = Much Improved | | | |

| **Supplementary Table 3.** List of Clinical Features used for Analysis | | |
| --- | --- | --- |
| **Group** | **Feature** | **Categories** |
| Demographics | Age Group | 18-39, 40-59, 60-79, 80+ |
|  | Race | Black, Asian, White, Unknown/Other |
|  | Ethnicity | Hispanic, Unknown/Other |
|  | Sex | Female, Male |
|  | Gender Identity | Female, Male, Non-Conforming, Trans-Female, Trans-Male, Not Disclosed |
|  | Sexual Orientation | Straight, Bisexual, Gay or Lesbian, Unknown |
| Socioeconomic Status | Marital Status | Single, Married, Widowed, Separated, Divorced, Unknown/Other |
|  | Religion | Christian, Muslim, Jewish, Buddhist, Hindu, Unspecified, Other |
|  | Preferred Language | English, Spanish, Not Collected, Other |
|  | County | New York, Bronx, Kings, Queens, Richmond, Brooklyn, Other |
|  | Housing Status | Domiciled, Undomiciled |
|  | Payor Financial Class | Medicaid, Medicare, Commercial/Managed, Other |
| Health-Related Characteristics | MyChart Status | Activated, Not Activated |
|  | Total Previous ED visits | 0-3 visits, 4-9 visits, 10-20 visits, 21-50 visits, 51-100 visits, 101+ visits |
|  | Smoking Status | Smoker, Non-smoker, Former-Smoker, Unknown |
| Clinical Encounter Details | Chief Complaint | GI, General, Substance Use Disorder, Glucose Metabolism, Ophthalmology, Surgical, GYN, Dental, ID, Dermatologic, GU, Psych, MSK, Cardiac, Respiratory, Trauma, Neuro, ENT, Gyn, Social, Other, Unspecified |
|  | Author Type | Registered Nurse, Physician Assistant, Resident, Physician |
|  | Acuity Level | Immediate (1), Emergent (2), Urgent (3), Less Urgent (4), non-urgent (5), Unspecified |
|  | Shift | 07:00-15:00, 15:00-23:00, 23:00-07:00 |

**Supplementary Figure 2.** Final Round Percentage Agreement and Cohen’s Kappa by Bias Category

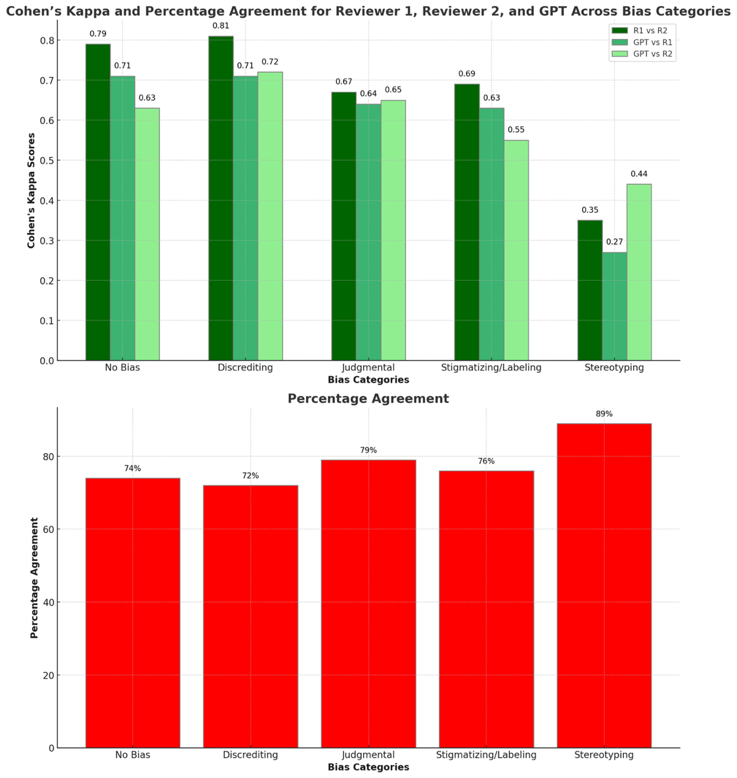

This figure illustrates the comparison between GPT's performance and human reviewers following the final round of prompt iteration, revealing important insights into bias identification in clinical documentation. Inter-rater reliability varied across bias categories. "Discrediting" showed highest reliability among human reviewers (Cohen's Kappa 0.81), while "Stereotyping" showed lowest (Kappa 0.35). Paradoxically, "Stereotyping" had 89% percentage agreement. Nevertheless, performance aligned with human reviewers, with slight deviations in "No Bias" and "Stigmatizing/Labeling" categories. "Discrediting," despite its high Kappa value, had the lowest percentage agreement (72%) between GPT and human reviewers.

| **Supplementary Table 4.** Comprehensive MSHS Patient Demographics | | | |
| --- | --- | --- | --- |
| **Group** | **Feature** | **Category** | **Number (%)** |
| Demographics  Socioeconomic Status | Age Group | 18-39 | 16,894 (33.8) |
|  |  | 40-59 | 14,552 (29.1) |
|  |  | 60-79 | 13,485 (27.0) |
|  |  | 80+ | 5,069 (10.1) |
|  | Race | Black | 15,711 (31.4) |
|  |  | Asian | 2,536 (5.1) |
|  |  | White | 12,673 (25.3) |
|  |  | Unknown/Other | 19,080 (38.2) |
|  | Ethnicity | Hispanic | 14,950 (29.9) |
|  |  | Unknown/Other | 35,050 (70.1) |
|  | Sex | Female | 27,019 (54.0) |
|  |  | Male | 22,977 (46.0) |
|  | Gender Identity | Female | 14,646 (29.3) |
|  |  | Male | 11,384 (22.8) |
|  |  | Non-Conforming | 116 (0.2) |
|  |  | Trans-Female | 70 (0.1) |
|  |  | Trans-Male | 18 (0.0) |
|  |  | Not Disclosed | 23,766 (47.5) |
|  | Sexual Orientation | Straight | 8,953 (17.9) |
|  |  | Bisexual | 307 (0.6) |
|  |  | Gay or Lesbian | 744 (1.5) |
|  |  | Unknown | 39,996 (80.0) |
|  | Marital Status | Single | 28,788 (57.6) |
|  |  | Married | 11,378 (22.8) |
|  |  | Widowed | 1,907 (3.8) |
|  |  | Separated | 3,613 (7.2) |
|  |  | Divorced | 1,684 (3.4) |
|  |  | Unknown/Other | 2,630 (5.3) |
|  | Religion | Christian | 22,452 (44.9) |
|  |  | Muslim | 2,246 (4.5) |
|  |  | Jewish | 1,996 (4.0) |
|  |  | Buddhist | 205 (0.4) |
|  |  | Hindu | 201 (0.4) |
|  |  | Unspecified | 5,260 (10.5) |
|  |  | Other | 4,597 (9.2) |
|  | Preferred Language | English | 43,729 (87.5) |
|  |  | Spanish | 4,581 (9.2) |
|  |  | Not Collected | 204 (0.4) |
|  |  | Other | 1,486 (3.0) |
|  | County | New York | 23,932 (47.9) |
|  |  | Bronx | 4,134 (8.3) |
|  |  | Kings | 7,721 (15.4) |
|  |  | Queens | 9,206 (18.4) |
|  |  | Richmond | 263 (0.5) |
|  |  | Brooklyn | 169 (0.3) |
|  |  | Other | 4,575 (9.2) |
|  | Housing Status | Domiciled | 47,697 (95.4) |
|  |  | Undomiciled | 2,303 (4.6) |
|  | Payor Financial Class | Medicaid | 18,531 (37.1) |
|  |  | Medicare | 14,618 (29.2) |
|  |  | Commercial/Managed | 12,787 (25.6) |
|  |  | Other | 4,064 (8.1) |
| Health-Related Characteristics | My Chart Status | Activated | 24,850 (49.7) |
|  |  | Not Activated | 25,150 (50.3) |
|  | Previous ED visits | 0-3 visits | 31,000 (62.0) |
|  |  | 4-9 visits | 10,172 (20.3) |
|  |  | 10-20 visits | 5,450 (10.9) |
|  |  | 21-50 visits | 2,593 (5.2) |
|  |  | 51-100 visits | 580 (1.2) |
|  |  | 101+ visits | 205 (0.4) |
|  | Smoking Status | Smoker | 6,922 (13.8) |
|  |  | Non-smoker | 24,386 (48.8) |
|  |  | Former-Smoker | 7,799 (15.6) |
|  |  | Unknown | 10,893 (21.8) |
| Clinical Encounter Details | Chief Complaint | GI | 6,863 (13.7) |
|  |  | General | 235 (0.5) |
|  |  | Substance Use Disorder | 553 (1.1) |
|  |  | Glucose Metabolism | 178 (0.4) |
|  |  | Ophthalmology | 852 (1.7) |
|  |  | Surgical | 171 (0.3) |
|  |  | GYN | 387 (0.8) |
|  |  | Dental | 427 (0.9) |
|  |  | ID | 815 (1.6) |
|  |  | Dermatologic | 575 (1.1) |
|  |  | GU | 660 (1.3) |
|  |  | Psych | 1,193 (2.4) |
|  |  | MSK | 8,030 (16.1) |
|  |  | Cardiac | 4,086 (8.2) |
|  |  | Respiratory | 3,924 (7.8) |
|  |  | Trauma | 2,727 (5.5) |
|  |  | Neuro | 4,934 (9.9) |
|  |  | ENT | 971 (1.9) |
|  |  | Gyn | 172 (0.3) |
|  |  | Social | 11 (0.0) |
|  |  | Other | 516 (1.0) |
|  |  | Unspecified | 11,720 (23.4) |
|  | Author Type | Registered Nurse | 26,130 (52.3) |
|  |  | Physician Assistant | 6,141 (12.3) |
|  |  | Resident | 4,291 (8.6) |
|  |  | Physician | 13,438 (26.9) |
|  | Acuity Level | Immediate (1) | 436 (0.9) |
|  |  | Emergent (2) | 11,212 (22.4) |
|  |  | Urgent (3) | 29,211 (58.4) |
|  |  | Less Urgent (4) | 8,219 (16.4) |
|  |  | Non-Urgent (5) | 622 (1.2) |
|  |  | Unspecified | 300 (0.6) |
|  | Shift | 07:00-15:00 | 21,795 (43.6) |
|  |  | 15:00-23:00 | 20,123 (40.2) |
|  |  | 23:00-07:00 | 8,082 (16.2) |

**Supplementary Figure 3.** Distribution of Bias Subtypes in MSHS Patient Notes

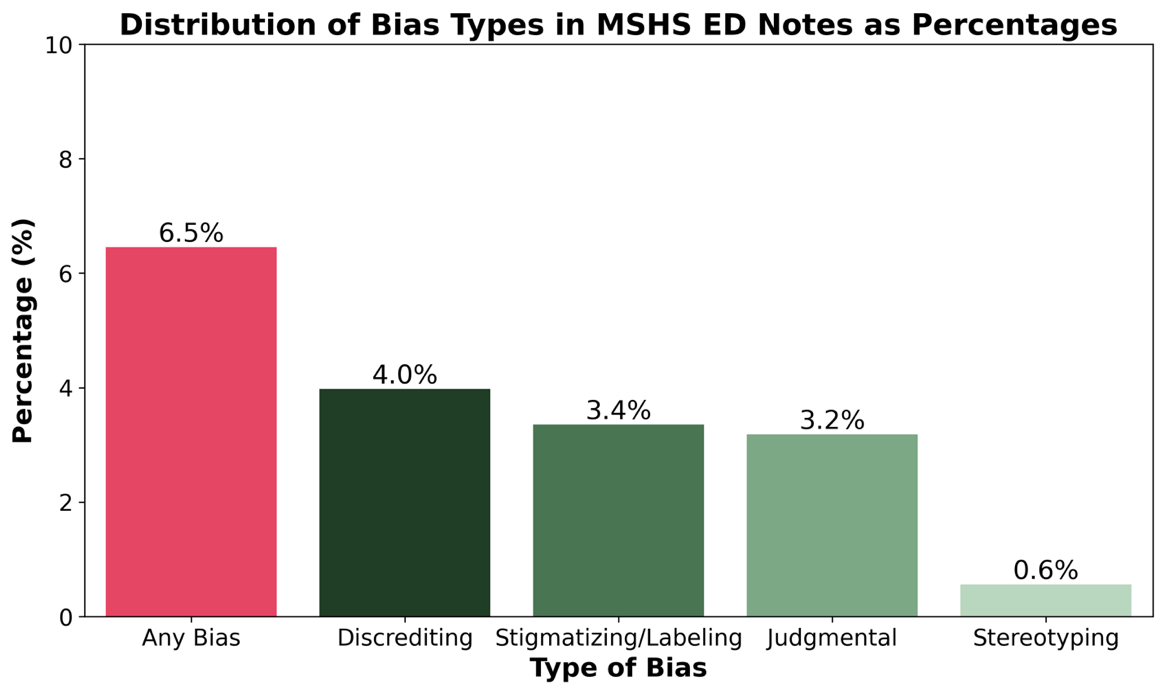

| **Supplementary Table 5.** Examples of Negative Descriptors Categories in the EMR with GPT Revision | |
| --- | --- |
|  | GPT Revision |
| Clinical Note: Unwitnessed fall but patient claimed "he tripped and fell" accompanied by HHA OOB with assistance only. At baseline with laceration at the back of the head | Patient reported that he tripped and fell. |
| Example of Judgmental Bias in EMR | GPT Revision |
| Clinical Note: Patient states he fell and injured his hip last week and has been snorting and injecting heroin to cope with pain. Last heroin use was yesterday. | Patient reports using heroin last week for self-medication of pain following a fall. |
| Example of Stigmatizing/Labeling Bias in EMR | GPT Revision |
| Clinical Note: Patient is c/o pain. Pain meds is given as order... patient is being disrespectful yelling and cursing and stated we didn't know what we were doing and wanted to report told patient to go ahead he has that right to do so. | Patient expressed concern loudly and seemed upset. |
| Example of Stereotyping Bias in EMR | GPT Revision |
| Clinical Note: 62M w/hx Afib (on eliquis), CVA, hyperthyroidism, etoh use disorder, htn, BIBEMS after someone called 911 as he was slumped on bench… Pt sleeping, opens eyes to verbal stimuli but turns away from provider and asks to be left alone…. No signs of head trauma, MAE. C/f acute alcohol intoxication vs. shelter seeking behavior. | Ruling out reasons for presentation, including potential alcohol intoxication or social needs, among others |

| **Supplementary Table 6.** Comprehensive MSHS Univariate Analysis | | | | | |
| --- | --- | --- | --- | --- | --- |
| **Group** | **Feature** | **Category** | **Bias** | **Bias Percentage 95% CI** | **P-value** |
| Demographics  Socioeconomic Status | Age Group | 18-39 | 1208 (7.2%) | (6.8, 7.5) | <0.001 |
|  |  | 40-59 | 1089 (7.5%) | (7.0, 7.9) |  |
|  |  | 60-79 | 706 (5.2%) | (4.9, 5.6) |  |
|  |  | 80+ | 226 (4.5%) | (3.9, 5.0) |  |
|  | Race | Black | 1298 (8.3%) | (7.8, 8.7) | <0.001 |
|  |  | Asian | 87 (3.4%) | (2.7, 4.1) |  |
|  |  | White | 755 (6.0%) | (5.6, 6.4) |  |
|  |  | Unknown/Other | 1089 (5.7%) | (5.4, 6.0) |  |
|  | Ethnicity | Hispanic | 823 (5.5%) | (5.1, 5.9) | <0.001 |
|  |  | Unknown/Other | 2406 (6.9%) | (6.6, 7.1) |  |
|  | Sex | Female | 1242 (4.6%) | (4.4, 4.9) | <0.001 |
|  |  | Male | 1987 (8.7%) | (8.3, 9.0) |  |
|  | Gender Identity | Female | 701 (4.8%) | (4.4, 5.1) | <0.001 |
|  |  | Male | 966 (8.5%) | (8.0, 9.0) |  |
|  |  | Non-Conforming | 17 (14.7%) | (8.2, 21.1) |  |
|  |  | Trans-Female | 11 (15.7%) | (7.2, 24.2) |  |
|  |  | Trans-Male | 3 (16.7%) | (-0.6, 33.9) |  |
|  |  | Not Disclosed | 1531 (6.4%) | (6.1, 6.8) |  |
|  | Sexual Orientation | Straight | 506 (5.7%) | (5.2, 6.1) | 0.001 |
|  |  | Bisexual | 29 (9.4%) | (6.2, 12.7) |  |
|  |  | Gay or Lesbian | 53 (7.1%) | (5.3, 9.0) |  |
|  |  | Unknown | 2641 (6.6%) | (6.4, 6.8) |  |
|  | Marital Status | Single | 2378 (8.3%) | (7.9, 8.6) | <0.001 |
|  |  | Married | 327 (2.9%) | (2.6, 3.2) |  |
|  |  | Widowed | 74 (3.9%) | (3.0, 4.8) |  |
|  |  | Separated | 199 (5.5%) | (4.8, 6.2) |  |
|  |  | Divorced | 86 (5.1%) | (4.1, 6.2) |  |
|  |  | Unknown/Other | 165 (6.3%) | (5.3, 7.2) |  |
|  | Religion | Christian | 1223 (5.5%) | (5.2, 5.8) | <0.001 |
|  |  | Muslim | 95 (4.2%) | (3.4, 5.1) |  |
|  |  | Jewish | 70 (3.5%) | (2.7, 4.3) |  |
|  |  | Buddhist | 10 (4.9%) | (1.9, 7.8) |  |
|  |  | Hindu | 5 (2.5%) | (0.3, 4.6) |  |
|  |  | Unspecified | 345 (6.6%) | (5.9, 7.2) |  |
|  |  | Other | 342 (7.4%) | (6.7, 8.2) |  |
|  | Preferred Language | English | 2988 (6.8%) | (6.6, 7.1) | <0.001 |
|  |  | Spanish | 181 (4.0%) | (3.4, 4.5) |  |
|  |  | Not Collected | 17 (8.3%) | (4.5, 12.1) |  |
|  |  | Other | 43 (2.9%) | (2.0, 3.7) |  |
|  | County | New York | 1886 (7.9%) | (7.5, 8.2) | <0.001 |
|  |  | Bronx | 300 (7.3%) | (6.5, 8.0) |  |
|  |  | Kings | 390 (5.0%) | (4.6, 5.5) |  |
|  |  | Queens | 355 (3.9%) | (3.5, 4.3) |  |
|  |  | Richmond | 29 (11.0%) | (7.2, 14.8) |  |
|  |  | Brooklyn | 7 (4.1%) | (1.1, 7.1) |  |
|  |  | Other | 262 (5.7%) | (5.1, 6.4) |  |
|  | Housing Status | Domiciled | 2607 (5.5%) | (5.3, 5.7) | <0.001 |
|  |  | Undomiciled | 622 (27.0%) | (25.2, 28.8) |  |
|  | Payor Financial Class | Medicaid | 1643 (8.9%) | (8.5, 9.3) | <0.001 |
|  |  | Medicare | 759 (5.2%) | (4.8, 5.6) |  |
|  |  | Commercial/  Managed | 427 (3.3%) | (3.0, 3.7) |  |
|  |  | Other | 400 (9.8%) | (8.9, 10.8) |  |
| Health-Related Characteristics | My Chart Status | Activated | 1080 (4.3%) | (4.1, 4.6) | <0.001 |
|  |  | Not Activated | 2149 (8.5%) | (8.2, 8.9) |  |
|  | Previous ED Visits | 0-3 visits | 1624 (5.2%) | (5.0, 5.5) | <0.001 |
|  |  | 4-9 visits | 703 (6.9%) | (6.4, 7.4) |  |
|  |  | 10-20 visits | 477 (8.8%) | (8.0, 9.5) |  |
|  |  | 21-50 visits | 289 (11.2%) | (9.9, 12.4) |  |
|  |  | 51-100 visits | 93 (16.0%) | (13.0, 19.0) |  |
|  |  | 101+ visits | 43 (21.0%) | (15.4, 26.6) |  |
|  | Smoking Status | Smoker | 956 (13.8%) | (13.0, 14.6) | <0.001 |
|  |  | Non-smoker | 1011 (4.2%) | (3.9, 4.4) |  |
|  |  | Former-Smoker | 345 (4.4%) | (4.0, 4.9) |  |
|  |  | Unknown | 917 (8.4%) | (7.9, 8.9) |  |
| Clinical Encounter Details | Chief Complaint | GI | 320 (4.7%) | (4.2, 5.2) | <0.001 |
|  |  | General | 6 (2.5%) | (0.5, 4.6) |  |
|  |  | Substance Use Disorder | 178 (32.2%) | (28.3, 36.1) |  |
|  |  | Glucose Metabolism | 30 (16.9%) | (11.4, 22.4) |  |
|  |  | Ophthalmology | 27 (3.2%) | (2.0, 4.4) |  |
|  |  | Surgical | 2 (1.2%) | (-0.4, 2.8) |  |
|  |  | GYN | 15 (3.9%) | (2.0, 5.8) |  |
|  |  | Dental | 17 (4.0%) | (2.1, 5.8) |  |
|  |  | ID | 23 (2.8%) | (1.7, 4.0) |  |
|  |  | Dermatologic | 31 (5.4%) | (3.5, 7.2) |  |
|  |  | GU | 22 (3.3%) | (2.0, 4.7) |  |
|  |  | Psych | 298 (25.0%) | (22.5, 27.4) |  |
|  |  | MSK | 379 (4.7%) | (4.3, 5.2) |  |
|  |  | Cardiac | 188 (4.6%) | (4.0, 5.2) |  |
|  |  | Respiratory | 159 (4.0%) | (3.4, 4.7) |  |
|  |  | Trauma | 128 (4.7%) | (3.9, 5.5) |  |
|  |  | Neuro | 370 (7.5%) | (6.8, 8.2) |  |
|  |  | ENT | 22 (2.3%) | (1.3, 3.2) |  |
|  |  | Gyn | 7 (4.1%) | (1.1, 7.0) |  |
|  |  | Social | 1 (9.1%) | (-7.9, 26.1) |  |
|  |  | Other | 18 (3.5%) | (1.9, 5.1) |  |
|  |  | Unspecified | 988 (8.4%) | (7.9, 8.9) |  |
|  | Author Type | Registered Nurse | 1373 (5.2%) | (5.0, 5.5) | <0.001 |
|  |  | Physician Assistant | 245 (4.0%) | (3.5, 4.5) |  |
|  |  | Resident | 343 (8.0%) | (7.2, 8.8) |  |
|  |  | Physician | 1268 (9.4%) | (9.0, 9.9) |  |
|  | Acuity Level | Immediate (1) | 28 (6.4%) | (4.1, 8.7) | <0.001 |
|  |  | Emergent (2) | 810 (7.2%) | (6.7, 7.7) |  |
|  |  | Urgent (3) | 1992 (6.8%) | (6.5, 7.1) |  |
|  |  | Less Urgent (4) | 346 (4.2%) | (3.8, 4.6) |  |
|  |  | Non-Urgent (5) | 25 (4.0%) | (2.5, 5.6) |  |
|  |  | Unspecified | 28 (9.3%) | (6.0, 12.6) |  |
|  | Shift | 07:00-15:00 | 1155 (5.3%) | (5.0, 5.6) | <0.001 |
|  |  | 15:00-23:00 | 1295 (6.4%) | (6.1, 6.8) |  |
|  |  | 23:00-07:00 | 779 (9.6%) | (9.0, 10.3) |  |

**Supplementary Figure 4.** Distribution of Bias Subtypes in MIMIC-IV Patient Notes
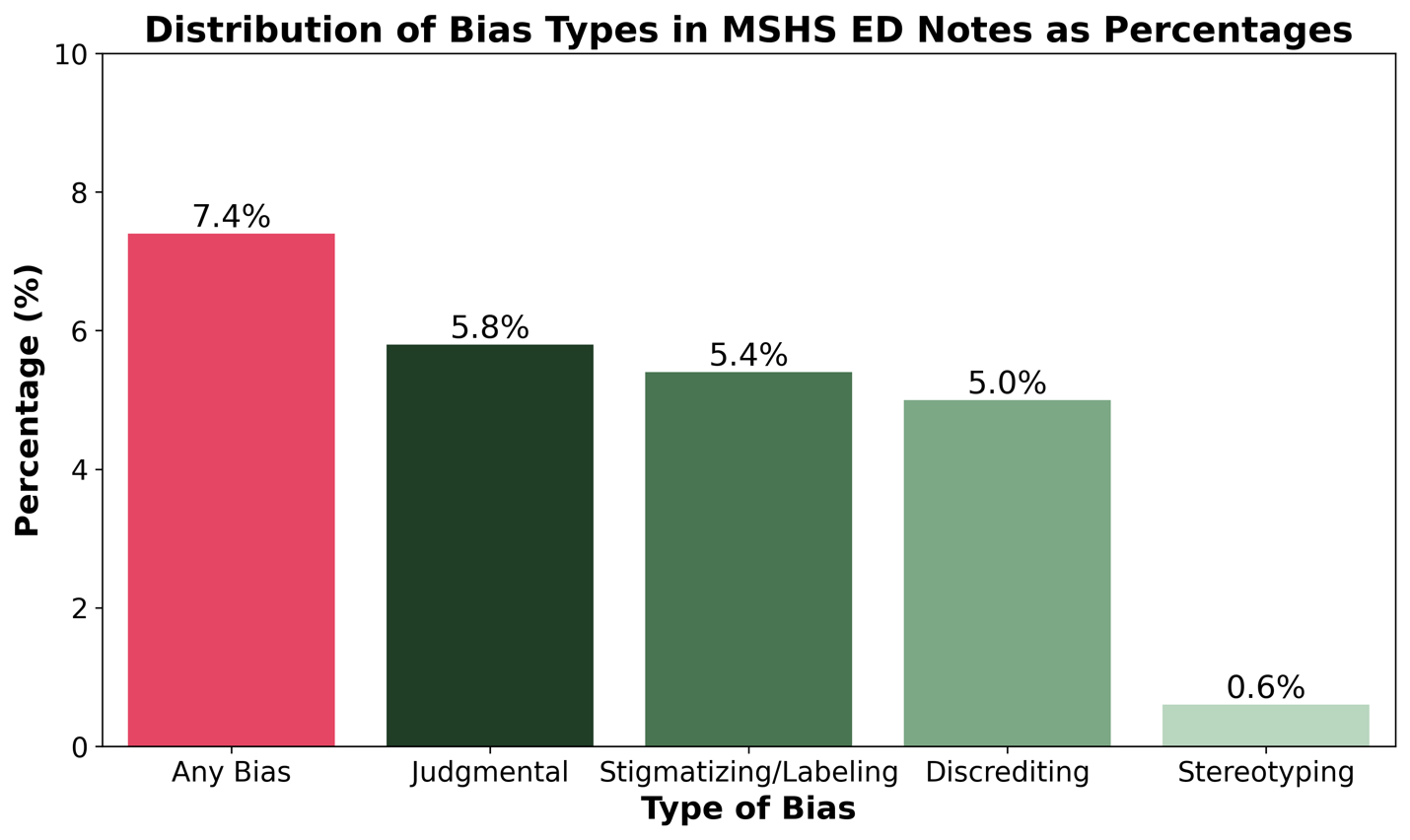

**REFERENCES.**

6.	Gulamali FF, Sawant AS, Liharska L, et al. An AI-Guided Data Centric Strategy to Detect and Mitigate Biases in Healthcare Datasets.

7..	Wei X, Cui X, Cheng N, et al. ChatIE: Zero-Shot Information Extraction via Chatting with ChatGPT. Published online February 20, 2023. <http://arxiv.org/abs/2302.10205>

8..	Liu Z, Huang Y, Yu X, et al. DeID-GPT: Zero-shot Medical Text De-Identification by GPT-4. Published online March 20, 2023. <http://arxiv.org/abs/2303.11032>
